# Humoral immune response of patients infected by earlier lineages of SARS-CoV-2 neutralizes wild types of the most prevalent variants in Brazil

**DOI:** 10.1101/2022.01.24.22269379

**Authors:** Alex Pauvolid-Corrêa, Braulia Costa Caetano, Ana Beatriz Machado, Mia Araújo Ferreira, Natalia Valente, Thayssa Keren Neves, Kim Geraldo, Fernando Motta, Valdiléa Gonçalves Veloso dos Santos, Beatriz Grinsztejn, Marilda Mendonça Siqueira, Paola Cristina Resende

## Abstract

In the present study, serum samples of 20 hospitalized COVID-19 patients from Brazil who were infected by the earlier SARS-CoV-2 lineages B.1.1.28 and B.1.1.33, and by the variant of concern (VOC) Gamma (P.1) were tested by plaque reduction neutralization test (PRNT_90_) with wild isolates of a panel of SARS-CoV-2 lineages, including B.1, Zeta, N.10, and the VOCs Gamma, Alpha, and Delta that emerged in different timeframes of the pandemic. The main objectives of the present study were to evaluate if serum of COVID-19 patients infected by earlier lineages of SARS-CoV-2 were capable to neutralize recently emerged VOCs, and if PRNT_90_ is a reliable serologic method to distinguish infections caused by different SARS-CoV-2 lineages. Overall, sera collected from the day of admittance to the hospital to 21 days after diagnostic of patients infected by the two earlier lineages B.1.1.28 and B.1.1.33 presented neutralizing capacity for all challenged VOCs, including Gamma and Delta, that were the most prevalent VOCs in Brazil. Among all variants tested, Delta and N.10 presented the lowest mean of neutralizing antibody titers, and B.1.1.7, presented the highest titers. Four patients infected with Gamma, that emerged in December 2020, presented neutralizing antibodies for B.1, B.1.1.33 and B.1.1.28, its ancestor lineage. All of them had neutralizing antibodies under the level of detection for the VOC Delta. Interestingly, patients infected by B.1.1.28 presented very similar mean of neutralizing antibody titers for both B.1.1.33 and B.1.1.28. Findings presented here indicate that most patients infected in early stages of COVID-19 pandemic presented neutralizing antibodies up to 21 days after diagnostic capable to neutralize wild types of all recently emerged VOCs in Brazil, and that the PRNT_90_ it is not a reliable serologic method to distinguish natural infections caused by different SARS-CoV-2 lineages.

## Introduction

The ongoing pandemic of coronavirus disease 2019 (COVID-19) is a major healthcare threat worldwide. While viral RNA-based testing for acute infection of severe acute respiratory syndrome coronavirus 2 (SARS-CoV-2) is the current standard, surveying antibodies is necessary not only to determine the past exposure, but also to identify donors with high-neutralizing titers for convalescent plasma for therapy [1]. Despite the relationship between humoral response and clinical protection from SARS-CoV-2 infection remains not fully understood, some studies have confirmed neutralizing antibodies as an immune correlate of protection [2]. The humoral immune response can block infection through neutralizing antibodies, which bind the virus in a manner that prevents host cell infection [3]. The host humoral response against SARS-CoV-2, including IgA, IgM, and IgG response, has been examined mostly by ELISA-based assays using recombinant viral nucleocapsid protein or pseudovirus-based neutralization assays [4]. Coronavirus infections typically induce neutralizing antibody responses, and virus neutralization assays performed on cell cultures, as plaque reduction neutralization test (PRNT), are considered as gold standard for serological testing and determining immune protection [1]. Although antiviral T and B cell memory certainly contribute some degree of protection, strong evidence of a protective role for neutralizing serum antibodies exists [5]. Neutralizing antibody levels are highly predictive of immune protection from symptomatic SARS-CoV-2 infection [2].

Since the pandemic began in China in December 2019, thousands of SARS-CoV-2 lineages have emerged worldwide [6]. The variants that presented increased transmissibility, virulence, and decreased response to available diagnostics, vaccines, and therapeutics were defined by the World Health Organization as variant of concern (VOC) [7]. Following the upsurge of variants, several reinfection cases started to be reported worldwide rising questions about the efficiency of humoral response mounted after primary infections to prevent a secondary infection by SARS-CoV-2 [8]. In fact, some recent studies have experimentally demonstrated that monoclonal antibodies, convalescent plasma of individuals infected by earlier lineages, and serum collected from vaccinees have a reduced neutralizing capacity when challenged with the recently emerged VOCs of SARS-CoV-2 [9–11].

The cross-reactivity of humoral responses among all different lineages of SARS-CoV-2 including VOCs, as well as the potential use of specific serological methods as PRNT to distinguish lineage infections remain unclear. Several lineages of SARS-CoV-2 have been reported in Brazil by the consortium COVID-19 Fiocruz Genomics Surveillance Network of the Brazilian Ministry of Health (http://www.genomahcov.fiocruz.br/dashboard/). Among the most important lineages detected in the country were the VOCs Gamma, Delta and Alpha, as well as the Zeta, B.1, B.1.1.28, B.1.1.33 and N.10. The main objectives of the present study were to assess the capacity of sera of COVID-19 patients infected by earlier lineages of SARS-CoV-2 to neutralize the two most prevalent VOCs in Brazil, and to evaluate if PRNT_90_ is a reliable serologic method to distinguish infections caused by different SARS-CoV-2 lineages.

## Methods

### Case description

Swab and serum samples were periodically collected from hospitalized COVID-19 patients that were admitted to the COVID-19 Hospital of the National Institute of Infectious Diseases, Rio de Janeiro. Samples were weekly collected from the day of admittance to up 21 days after diagnostic. The index cases were patients aged 18 years and older, with SARS-CoV-2 infection confirmed by real-time reverse transcription polymerase chain reaction (qRT-PCR) in respiratory samples, which consisted of a combination of two nasopharyngeal swabs and one oropharyngeal swab collected in 3 mL of viral transportation medium (VTM).

### qRT-PCR for SARS-CoV-2

The VTM of respiratory samples were submitted to RNA extraction using Chemagic Viral DNA/RNA 300 kit H96 in a Janus G3 automated workstation (Perkin Elmer, www.chemagen.com). SARS-CoV-2 was detected by qRT-PCR assays targeting the viral gene E. Reactions were performed using the Kit Molecular SARS-CoV-2 (E/RP) (Bio-Manguinhos/IBMP, Brazil).

### Whole-genome sequencing

Positive samples eligible for whole-genome sequencing had RNA extracted using the QIAamp Viral RNA Mini Kit (QIAGEN) or using Chemagic Viral DNA/RNA 300 kit H96 in a Janus G3 automated workstation (Perkin Elmer, www.chemagen.com). The SARS-CoV-2 genomes were recovered by amplification of long segments (2kb), according to a protocol developed by COVID-19 Fiocruz Genomic Surveillance Network to recover high-quality genomes (P.C. Resende, unpub. data, doi.org/10.1101/2020.04.30.069039). Segment libraries were then sequenced in Illumina MiSeq. The FASTQ reads obtained were imported into the CLC Genomics Workbench version 20.0.4 (QIAGEN), trimmed, and mapped against the reference sequence hCoV-19/Wuhan/WIV04/2019 (GISAID access number EPI_ISL_402124) to obtain the final genome consensus. The SARS-CoV-2 lineage characterization was performed by Pango Network [12]. All genomic and epidemiological data associated were uploaded at the EpiCoV database in the GISAID (www.gisaid.org).

### Isolation of SARS-CoV-2 reference lineages used for PRNT

As part of the Brazil’s Surveillance Network for Respiratory Illnesses, respiratory samples collected in sentinel units located in different states of the country are routinely sent to the Reference Laboratory. Eligible samples that tested positive for different lineages of SARS-CoV-2 were submitted to virus isolation in Vero E6 or Vero CCL-81 cells at a Biosafety level 3 Laboratory, as previously described [13]. Briefly, 200 μL of each respiratory sample were inoculated in cell cultures, which were then inspected daily for cytopathic effect (CPE), up to four days. For each sample, isolation was attempted in a maximum of three consecutive blind passages. Overall, in positive cultures, CPE started on second day post-infection and viral harvest was performed at the fourth day post-infection. When CPE was observed, culture supernatants were aliquoted in working stocks and an aliquot submitted to qRT-PCR followed by nucleotide sequencing for lineage confirmation. Once confirmed, the consensus sequences were deposited at the EpiCoV data base on GlSAID (www.gisaid.org), and one representative of each SARS-CoV-2 lineage sequentially titrated by lysis plaque assay, to form a lineage bank with reference isolates.

### PRNT

Subsets of serum samples of hospitalized patients that were positive for different lineages of SARS-CoV-2 were selected for PRNT. For the patients who had several blood samples collected during hospitalization, the last samples were preferred. Briefly, sera were heat-inactivated at 56°C for 30 minutes do inactivate the complement system. Inactivated serum samples were submitted to PRNT_90_ in Vero CCL-81 cells in duplicate at serial 2-fold dilutions to determine 90% endpoint titers against five infectious SARS-CoV-2 reference lineages isolates. The panel of reference isolates used for PRNT included the lineages B.1 (hCoV-19/Brazil/RJ-FIOCRUZ-314/2020, GISAID accession number EPI_ISL_414045), B.1.1.28 (hCoV-19/Brazil/AL-FIOCRUZ-33444-1P/2020, EPI_ISL_2645638), B.1.1.33 (hCoV-19/Brazil/RJ-FIOCRUZ-20136-1P/2020, EPI_ISL_1181430), Zeta (hCoV-19/Brazil/PB-FIOCRUZ-33096-1P/2020, EPI_ISL_1402429), N.10 (hCoV-19/Brazil/MA-FIOCRUZ-6871-1P/2021, EPI_ISL_3828018), and the VOCs Gamma (hCoV-19/Brazil/AM-FIOCRUZ-3521-1P/2021, EPI_ISL_1402431), Alpha (hCoV-19/Brazil/RJ-FIOCRUZ-2624-1P/2021, EPI_ISL_1402430) and Delta (hCoV-19/Brazil/MA-FIOCRUZ-25688-2P/2021, EPI_ISL_2645417). Neutralizing antibodies were quantified, and serum samples were considered seropositive when at least 1:10 dilution reduced ≥90% of the formation of SARS-CoV-2 plaque forming units (PFU), as previously reported [14].

## Ethics

This study was approved by the Ethics Committee of the Fundação Oswaldo Cruz (CAAE 68118417.6.0000.5248), and the Ethics Committee of the National Institute of Infectious Diseases (CAAE 32449420.4.1001.5262). All patients entering the study were required to read and sign an informed consent form.

## Results

From a population of patients recruited from May 2020 to August 2021, we were able to obtain a panel of sera of individuals infected by SARS-CoV-2 lineages B.1.1.28, B.1.1.33, the two most common lineages in early pandemic [15] and Gamma, the first VOC to become prevalent in Brazil [16]. From a total of 20 serum samples evaluated, five were from patients positive for B.1.1.28, four from patients that tested positive for Gamma, and 11 were positive for B.1.1.33 (Table). Among five patients infected by B.1.1.28, highest neutralizing antibody titers were observed for B.1.1.28, followed by Alpha, B.1.1.33, B.1, Gamma, Zeta, N.10 and Delta (Figure). Among patients infected by B.1.1.33, similar profile was observed. Neutralizing antibody titer mean was higher for B.1.1.33, followed by Alpha, B.1, B.1.1.28, Zeta, Gamma, and with equally reduced neutralizing antibodies for Delta and N.10 (Figure). From four patients infected by Gamma, highest PRNT_90_ titers were observed for Gamma, followed by Alpha, B.1.1.33, N.10, B.1.1.28 and Zeta, and with neutralizing antibodies under the level of detection for Delta in all individuals (Table).

**Table:**
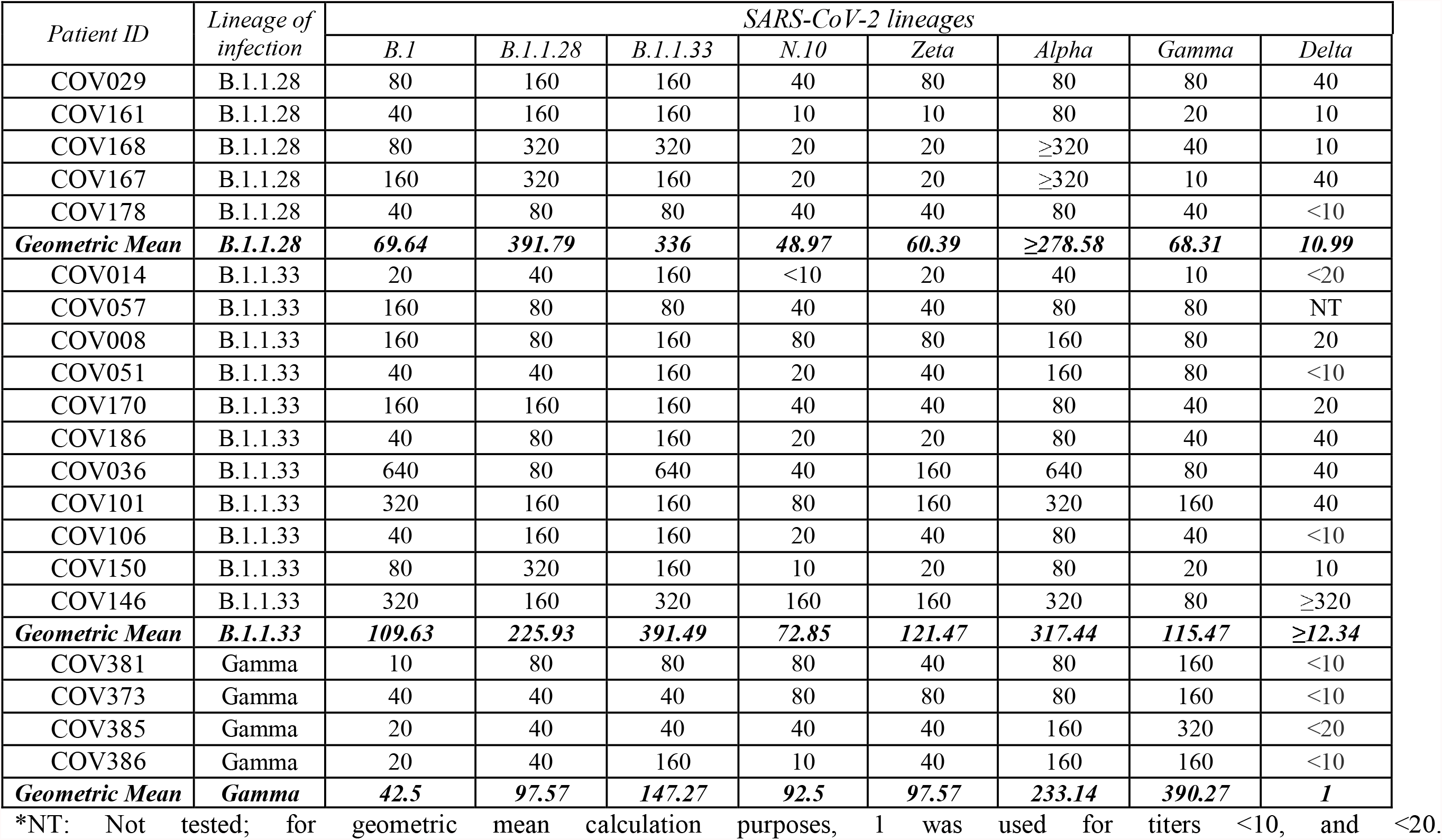
PRNT_90_ titers of convalescent sera of COVID-19 patients for different SARS-CoV-2 lineages

| Patient ID | Lineage of infection | SARS-CoV-2 lineages |  |  |  |  |  |  |  |
| --- | --- | --- | --- | --- | --- | --- | --- | --- | --- |
|  |  | *B.1* | *B.1.1.28* | *B.1.1.33* | *N.10* | *Zeta* | *Alpha* | *Gamma* | *Delta* |
| COV029 | B.1.1.28 | 80 | 160 | 160 | 40 | 80 | 80 | 80 | 40 |
| COV161 | B.1.1.28 | 40 | 160 | 160 | 10 | 10 | 80 | 20 | 10 |
| COV168 | B.1.1.28 | 80 | 320 | 320 | 20 | 20 | ≥320 | 40 | 10 |
| COV167 | B.1.1.28 | 160 | 320 | 160 | 20 | 20 | ≥320 | 10 | 40 |
| COV178 | B.1.1.28 | 40 | 80 | 80 | 40 | 40 | 80 | 40 | <10 |
| **Geometric Mean** | **B.1.1.28** | **69.64** | **391.79** | **336** | **48.97** | **60.39** | **≥278.58** | **68.31** | **10.99** |
| COV014 | B.1.1.33 | 20 | 40 | 160 | <10 | 20 | 40 | 10 | <20 |
| COV057 | B.1.1.33 | 160 | 80 | 80 | 40 | 40 | 80 | 80 | NT |
| COV008 | B.1.1.33 | 160 | 80 | 160 | 80 | 80 | 160 | 80 | 20 |
| COV051 | B.1.1.33 | 40 | 40 | 160 | 20 | 40 | 160 | 80 | <10 |
| COV170 | B.1.1.33 | 160 | 160 | 160 | 40 | 40 | 80 | 40 | 20 |
| COV186 | B.1.1.33 | 40 | 80 | 160 | 20 | 20 | 80 | 40 | 40 |
| COV036 | B.1.1.33 | 640 | 80 | 640 | 40 | 160 | 640 | 80 | 40 |
| COV101 | B.1.1.33 | 320 | 160 | 160 | 80 | 160 | 320 | 160 | 40 |
| COV106 | B.1.1.33 | 40 | 160 | 160 | 20 | 40 | 80 | 40 | <10 |
| COV150 | B.1.1.33 | 80 | 320 | 160 | 10 | 20 | 80 | 20 | 10 |
| COV146 | B.1.1.33 | 320 | 160 | 320 | 160 | 160 | 320 | 80 | ≥320 |
| **Geometric Mean** | **B.1.1.33** | **109.63** | **225.93** | **391.49** | **72.85** | **121.47** | **317.44** | **115.47** | **≥12.34** |
| COV381 | Gamma | 10 | 80 | 80 | 80 | 40 | 80 | 160 | <10 |
| COV373 | Gamma | 40 | 40 | 40 | 80 | 80 | 80 | 160 | <10 |
| COV385 | Gamma | 20 | 40 | 40 | 40 | 40 | 160 | 320 | <20 |
| COV386 | Gamma | 20 | 40 | 160 | 10 | 40 | 160 | 160 | <10 |
| **Geometric Mean** | **Gamma** | **42.5** | **97.57** | **147.27** | **92.5** | **97.57** | **233.14** | **390.27** | **1** |

\*NT: Not tested; for geometric mean calculation purposes, 1 was used for titers <10, and <20.

**Figure:**
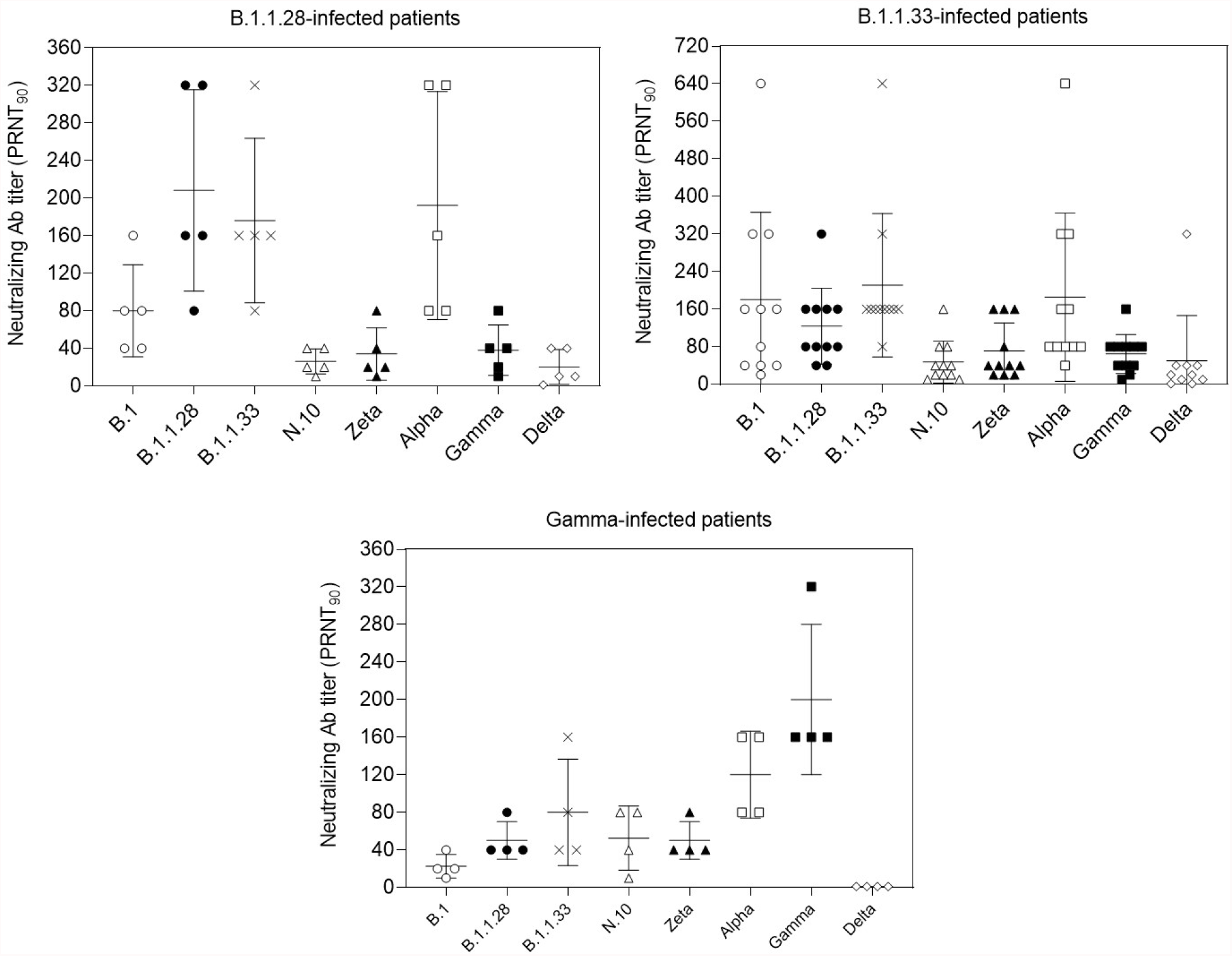
Anti-SARS-CoV-2 neutralizing antibodies titers in serum samples form COVID-19 patients admitted to a reference hospital in Rio de Janeiro, Brazil. Samples were obtained from patients infected with the SARS-CoV-2 lineages B.1.1.28 (top left), B.1.1.33 (top right) or Gamma (P.1, bottom panel). Samples were heat-inactivated, submitted to limiting dilution and then tested against different viral isolates (x axis of each panel) belonging to early pandemic lineages (B.1; B.1.1.28 and B.1.1.33), variants of interest (N.10 and P.2) and variants of concern (Alpha, Gamma, and Delta).

## Discussion

The relationship between humoral response to SARS-CoV-2 antigens, especially the spike protein, and clinical protection from COVID-19 remains not fully understood, although some studies have confirmed neutralizing antibodies as an immune correlate of protection [2]. Likewise, it is still not completely understood to what extent the mutations in viral antigens contribute to virus evasion from neutralizing antibodies, and how significant this escape mechanism could be for the general effectiveness of the protective response, especially in the context of vaccinations, reinfections, and the evolution of the pandemic. It has been shown that mutations in the ACE2 binding site of variants result in an increased affinity for the receptor ACE2, and that changes outside the receptor-binding domain also impact neutralization [17]. In this scenario, we primarily investigated the neutralization profile of sera from patients infected by the most prevalent lineages in early pandemic in Brazil including B.1.1.33 and B.1.1.28, with the recently emerged VOCs including Gamma, Alpha, and Delta. Additionally, we investigated if Gamma-induced antibodies were capable to neutralize the early isolates and the most recently emerged VOC, Delta. We observed that, despite the variation in antibody levels, most serum samples of patients infected by early isolates presented some level of neutralizing activity against all VOCs. The reverse situation, in which serum samples from individuals infected with the Gamma lineage were challenged with early pandemic viral isolates, also produced detectable neutralization. However, when sera from Gamma-infected individuals were tested with other VOC Delta, the PRNT_90_ titer was below the limit of detection of the assay (Table).

Gamma, which emerged from the B.1.1.28 lineage, contains 17 amino acid substitutions, ten of which in the spike protein, including N501Y, E484K and K417T in the receptor-binding domain, five in the N-terminal domain, and the mutation H655Y near the furin cleavage site [18]. These mutations reduce the neutralization capacity of convalescent sera from individuals infected by early isolates [11], and findings presented here suggest these mutations perhaps interfere also with the efficiency of its neutralizing antibodies for other variants, as Delta.

Despite the small number of patients evaluated, the findings that Gamma-induced immune sera presented low or absent PRNT_90_ titers for Delta raises concern over the potential risk of reinfections, and consequent prolongation of the pandemic. Gamma was the most prevalent lineage of SARS-CoV-2 in Brazil for several months in a row, and now, people who were previously infected by Gamma are being exposed to Delta, which is currently the most prevalent VOC in the country [16]. In Brazil, Delta was first detected in May 2021, and by the end of August, 70% of the genomes of SARS-CoV-2 sequenced were Delta (www.genomahcov.fiocruz.br/dashboard/). As previously reported, this variant presents a higher replication fitness and decreased sensitivity to neutralizing antibodies mounted for early isolates, both contributing to higher transmissibility [19].

It is important to mention that the protective role of neutralizing antibodies does not seem to be directly linked to the titers in which they are found, but to timing and kinetics of their production, with studies suggesting a limited role of antibodies in predicting disease severity of the COVID-19, and that the earlier the presence of neutralizing antibodies after infection, the less severe is the disease outcome [20].

It is noteworthy that for most viruses there is no direct correlate of protection in humans, since the studies needed to establish such a correlate in humans are challenging. For instance, protective neutralizing antibody titers have been roughly estimated for yellow fever vaccine by challenge studies in nonhuman primates and hamster models [21, 22]. The threshold of protective neutralizing antibody titers for SARS-CoV-2 and its lineages has not been established. Therefore, lower neutralizing antibodies titers including the samples with titers under the limit of detection as <10, do not necessarily mean susceptibility to a hypothetical secondary challenge. Samples that presented titers below 10 were not tested in lower dilutions, and if titers ≤9 are protective for each one of the different lineages of SARS-CoV-2 tested remain unknown.

Despite concerns over the potential risk of infections caused by Delta variant in patients previously exposed to Gamma, it is noteworthy that the number of reinfections reported worldwide remains limited, and that reinfections caused by Delta variant after previous infection by another VOC, as Gamma, have not been reported yet. The continuous advance of the COVID-19 vaccination has reduced the pace of new infections worldwide and is also expected to mitigate the number of reinfections as well. It is important to mention though that the time Delta variant became dominant in Brazil coincides with the local increasing vaccination coverage, but because of the unequal pace of vaccination throughout the country and along the time, Brazil remains with vaccination coverage below desirable. By the beginning of October 2021, over 58% of the adult population had been fully vaccinated in Brazil [23]. Regardless the recent improvement, a recent study conducted in the United States found that if in the state of Florida, the coverage was 74% instead of 59,5%, vaccination would have averted 664,007 additional cases, and reduced hospital admissions by 61,327, and deaths by 16,235 [24].

Finally, cellular immune response is believed to play also an important role in the immune response [5]. Cellular immune response was not investigated in the present study, and only with a combined evaluation of both immune responses for a better understanding of the relationship between neutralizing antibody titers and susceptibility to secondary infections.

Besides being considered markers of immune protection, specific neutralizing antibodies have also been used to evaluate viral exposure, and ultimately, for diagnostic purposes. Assays designed to detect neutralization antibodies have been widely used for the diagnostic of different viral groups. The PRNT is the most specific and gold standard serological test for the differentiation of closely related flavivirus infections, as dengue and yellow fever viruses in convalescent serum samples [25]. Type-specific antibodies can be distinguished using the PRNT, and two or more flaviviruses are distinct from each other by quantitative serological criteria. Four-fold or greater differences between PRNT titers have been used as seropositivity criterion in heterologous reactions [25,26]. In the present study, despite some genetic and immunogenic differences observed among all lineages of SARS-CoV-2 worldwide, these viruses are closely related and for that reason a PRNT with a highly conservative threshold of 90% neutralization in ≥1:10 serum dilution was used as seropositivity criterion.

According to our findings, the four-fold or greater difference among PRNT titers was not a suitable seropositivity criterion to distinguish infections among the different lineages of SARS-CoV-2 tested. Convalescent serum from patients infected by B.1.1.28 presented a mean of PRNT titer very similar for B.1.1.28, B.1.1.33 and Alpha lineages, and the titer difference was less than four-fold greater than titers for B.1. Four-fold or greater titers for B.1.1.28 were observed only when compared to the titers of the variants N.10, Zeta and Delta.

The similar profile was observed for samples from patients infected by B.1.1.33. In this case, these patients presented a non-significant difference in PRNT titers between B.1.1.33, B.1 and Alpha. B.1.1.33-induced sera presented a mean of PRNT titer less than four-fold greater for B.1.1.33 when compared to B.1.1.28, Zeta and N.10. Four-fold or greater B.1.1.33 titers were observed only when compared to Gamma and Delta VOCs. Finally, for the patients previously infected by Gamma, PRNT titers for Gamma were more than four-fold higher for Gamma when compared to B.1 and B.1.1.28, Delta and Zeta. Differences were smaller than four-fold when compared to B.1.1.33, N.10, and Alpha. Of note, the mean of PRNT_90_ titers for the Alpha variant was the second highest in all the three groups of patients, which includes samples of patients infected by B.1.1.28 and B.1.1.33 that were collected before the upsurge of Alpha variant in the United Kingdom.

The difference in PRNT_90_ titers demonstrated here among all lineages indicate that the four-fold or greater criterion of seropositivity is not reliable to distinguish lineage infections. The same is truth if considering optionally two-fold or greater differences as criterion de seropositivity.

In conclusion, the humoral response mounted for more recently emerged variants after natural infections caused by earlier isolates and different lineages reported here, corroborates the importance of the international recommendation of a high vaccination coverage which has being instrumental to reduce severe cases and deaths, mitigate virus propagation, and the upsurge of new variants.

## Data Availability

All data produced in the present study are available upon reasonable request to the authors

## Acknowledgments

We gratefully acknowledge the patients and authors from the originating laboratories responsible for obtaining the specimens. We thank the funding support from General Laboratories Coordination of Brazilian Ministry of Health; the Coordination of Health Surveillance and Reference Laboratories of Oswaldo Cruz Foundation; Ministério da Ciência, Tecnologia, Inovações e Comunicações/Conselho Nacional de Desenvolvimento Científico e Tecnológico -CNPq/Ministério da Saúde–MS/FNDCT/SCTIE/Decit (grants no. 402457/2020-9 and 403276/2020-9); Inova Fiocruz/Fundação Oswaldo Cruz (grants no. VPPCB-007-FIO-18-2-30; FAPERJ: E26/210.196/2020). We also thank the support of the Fiocruz COVID-19 Genomic Surveillance Network (http://www.genomahcov.fiocruz.br), the Respiratory Viruses Genomic Surveillance Network of the General Laboratory Coordination, Brazilian Ministry of Health, Brazilian Central Laboratory States, the Multi-user Research Facility of Biosafety Level 3 Platform of the Oswaldo Cruz Institute and Fiocruz Diagnoses Unit Rio de Janeiro.

## Notes

### Competing Interest Statement

The authors have declared no competing interest.

### Funding Statement

This study was funded by: General Laboratories Coordination of Brazilian Ministry of Health; the Coordination of Health Surveillance and Reference Laboratories of Oswaldo Cruz Foundation; Ministerio da Ciencia, Tecnologia, Inovacoes e Comunicacoes/Conselho Nacional de Desenvolvimento Cientifico e Tecnologico CNPq/Ministerio da SaudeMS/FNDCT/SCTIE/Decit (grants no. 402457/2020-9 and 403276/2020-9); Inova Fiocruz/Fundacao Oswaldo Cruz (grants no. VPPCB-007-FIO-18-2-30; FAPERJ: E26/210.196/2020).

### Author Declarations

This study was approved by the Ethics Committee of the Fundacao Oswaldo Cruz (CAAE 68118417.6.0000.5248), and the Ethics Committee of the National Institute of Infectious Diseases (CAAE 32449420.4.1001.5262).

## References

1. Muruato AE, Fontes-Garfias CR, Ren P, Garcia-Blanco MA, Menachery VD, et al. Author Correction: A high-throughput neutralizing antibody assay for COVID-19 diagnosis and vaccine evaluation. Nat Commun 2021;12:4000.

2. Khoury DS, Cromer D, Reynaldi A, Schlub TE, Wheatley AK, et al. Neutralizing antibody levels are highly predictive of immune protection from symptomatic SARS-CoV-2 infection. Nat Med 2021;27:1205–1211.

3. VanBlargan LA, Goo L, Pierson TC. Deconstructing the Antiviral Neutralizing-Antibody Response: Implications for Vaccine Development and Immunity. Microbiol Mol Biol Rev 2016;80:989–1010.

4. Tang MS, Case JB, Franks CE, Chen RE, Anderson NW, et al. Association between SARS-CoV-2 Neutralizing Antibodies and Commercial Serological Assays. Clin Chem 2019;66:1538–1547.

5. Le Bert N, Tan AT, Kunasegaran K, Tham CYL, Hafezi M, et al. SARS-CoV-2-specific T cell immunity in cases of COVID-19 and SARS, and uninfected controls. Nature 2020;584:457–462.

6. Koyama T, Platt D, Parida L. Variant analysis of SARS-CoV-2 genomes. Bull World Health Organ 2020;98:495–504.

7. Konings F, Perkins MD, Kuhn JH, Pallen MJ, Alm EJ, et al. SARS-CoV-2 Variants of Interest and Concern naming scheme conducive for global discourse. Nat Microbiol 2021;6:821–823.

8. Babiker A, Marvil CE, Waggoner JJ, Collins MH, Piantadosi A. The Importance and Challenges of Identifying SARS-CoV-2 Reinfections. J Clin Microbiol;59. Epub ahead of print 19 March 2021. DOI: 10.1128/JCM.02769-20.

9. Vidal SJ, Collier AY, Yu J, McMahan K, Tostanoski LH, et al. Correlates of Neutralization against SARS-CoV-2 Variants of Concern by Early Pandemic Sera. J Virol;95. Epub ahead of print 2021. DOI: 10.1128/jvi.00404-21.

10. Liu C, Ginn HM, Dejnirattisai W, Supasa P, Wang B, et al. Reduced neutralization of SARS-CoV-2 B.1.617 by vaccine and convalescent serum. Cell;184. Epub ahead of print 2021. DOI: 10.1016/j.cell.2021.06.020.

11. Dejnirattisai W, Zhou D, Supasa P, Liu C, Mentzer AJ, et al. Antibody evasion by the P.1 strain of SARS-CoV-2. Cell;184. Epub ahead of print 2021. DOI: 10.1016/j.cell.2021.03.055.

12. O’Toole Á, Scher E, Underwood A, Jackson B, Hill V, et al. Assignment of epidemiological lineages in an emerging pandemic using the pangolin tool. Virus Evol 2021;7:1–9.

13. Basile K, McPhie K, Carter I, Alderson S, Rahman H, et al. Cell-based Culture Informs Infectivity and Safe De-Isolation Assessments in Patients with Coronavirus Disease 2019. Clin Infect Dis 2020;1–8.

14. Deshpande G, Sapkal G, Tilekar B, Yadav P, Gurav Y, et al. Neutralizing antibody responses to SARS-CoV-2 in COVID-19 patients. Indian J Med Res 2020;152:82.

15. Resende PC, Delatorre E, Gräf T, Mir D, Motta FC, et al. Evolutionary Dynamics and Dissemination Pattern of the SARS-CoV-2 Lineage B.1.1.33 During the Early Pandemic Phase in Brazil. Front Microbiol 2021;11:1–14.

16. Naveca FG, Nascimento V, de Souza VC, Corado A de L, Nascimento F, et al. COVID-19 in Amazonas, Brazil, was driven by the persistence of endemic lineages and P.1 emergence. Nat Med 2021;27:1230–1238.

17. Dejnirattisai W, Zhou D, Ginn HM, Duyvesteyn HME, Supasa P, et al. The antigenic anatomy of SARS-CoV-2 receptor binding domain. Cell;184. Epub ahead of print 2021. DOI: 10.1016/j.cell.2021.02.032.

18. Wang P, Casner RG, Nair MS, Wang M, Yu J, et al. Increased resistance of SARS-CoV-2 variant P.1 to antibody neutralization. Cell Host Microbe 2021;29:747–751.e4.

19. Mlcochova P, Kemp S, Dhar MS, Papa G, Meng B, et al. SARS-CoV-2 B.1.617.2 Delta variant replication and immune evasion. Nature. Epub ahead of print 2021. DOI: 10.1038/s41586-021-03944-y.

20. Ren L, Zhang L, Chang D, Wang JJ, Hu Y, et al. The kinetics of humoral response and its relationship with the disease severity in COVID-19. Commun Biol;3. Epub ahead of print 2020. DOI: 10.1038/s42003-020-01526-8.

21. Julander JG, Trent DW, Monath TP. Immune correlates of protection against yellow fever determined by passive immunization and challenge in the hamster model. Vaccine 2011;29:6008–6016.

22. Mason RA, Tauraso NM, Spertzel RO, Ginn RK. Yellow Fever Vaccine: Direct Challenge of Monkeys Given Graded Doses of 17D Vaccine. Appl Microbiol 1973;25:539–544.

23. Brazilian Ministry of Health. Vacinação COVID-19 - doses aplicadas. https://www.gov.br/saude/pt-br/vacinacao/ (2021).

24. Sah P, Moghadas SM, Vilches TN, Shoukat A, Singer BH, et al. Implications of suboptimal COVID-19 vaccination coverage in Florida and Texas. Lancet Infect Dis 2021;21:1493–1494.

25. Calisher CH, Karabatsos N, Dalrymple JM, Shope RE, Porterfield JS, et al. Antigenic Relationships between Flaviviruses as Determined by Cross-neutralization Tests with Polyclonal Antisera. J Gen Virol 1989;70:37–43.

26. Johnson BW, Kosoy O, Hunsperger E, Beltran M, Delorey M, et al. Evaluation of Chimeric Japanese Encephalitis and Dengue Viruses for Use in Diagnostic Plaque Reduction Neutralization Tests. Clin Vaccine Immunol 2009;16:1052–1059.

